# Analytical Centralization of Health Expenditure at the National Administrator of Health System Resources: Architecture, Data Quality, and Operational Performance of the ADRES Health System Analytics Platform, Colombia

**DOI:** 10.64898/2026.06.08.26355159

**Authors:** Daniel Alfonso Garavito Jiménez, David E. Bello Angulo, Lady Tatiana Mejía Lemus, Diana Chipatecua, Daniel Darío Fula, Santiago Perez-Rubiano, Félix León Martínez, Juan Camilo Bohórquez Pinzón

## Abstract

**Background:** Between 2024 and 2025, Colombia universalized the Electronic Health Invoice with embedded Individual Health Services Delivery Records (RIPS — Registro Individual de Prestación de Servicios de Salud) (FEV-RIPS) as the standard for financial and clinical data exchange across the health system. ADRES — the entity responsible for administering the resources of the General Social Security Health System (SGSSS) — faced the challenge of processing information from multiple heterogeneous sources generated by more than 55,000 healthcare providers of varying complexity. Health systems in high-income countries converge clinical-financial data in consolidated platforms with years of operation; Colombia started from a fragmented architecture with incompatible historical sources, no cross-database standardization, and a resource administrator with no centralized analytical infrastructure until 2023.

**Objective:** We describe the design, the technical challenges of integrating heterogeneous data, and the operational performance of the analytical infrastructure built by ADRES to centralize large-scale processing of Colombian health system information, and we derive transferable lessons for health system resource administrators in Latin America facing equivalent digitalization mandates.

**Methods:** Technical-descriptive report based on operational metrics from the ADRES Azure/Databricks environment during January–November 2025. We report indicators of data volume managed, processing speed, deployed computational capacity, concurrent use by functional group, and implemented governance structure. The architecture integrates secure data transfer with MinSalud via VPN, OneLake Fabric connectivity, automated processing of multiple formats (XML, relational tables, flat files), and a data lake with a medallion pattern (Bronze/Silver/Gold) and automated pipelines. Data quality challenges are characterized through structural inconsistencies across system sources, coding incompatibilities (municipalities, dates, diagnoses), format heterogeneities in unstructured data, and the absence of complete technical documentation.

**Results:** The platform manages 21 catalogs, 1,183 tables, and over 110,645 million stored records, with cumulative production exceeding 1 trillion processed records. It executes queries on 100 billion records in ten seconds, using clusters of up to 32 TB RAM and 4,096 vCPU. During September–October 2025, monthly query peaks reached 78,028, distributed across eleven institutional functional groups. Integrating heterogeneous sources required developing specific technical capabilities: Python/PySpark parsers for XML with variable node depth, institutional equivalence tables to homologate incompatible municipality codes between BDUA and service delivery records, cleaning routines for extreme dates used as null representations (1900-01-01, 9999-12-31), and transformation logic to build coherent longitudinal series bridging classic RIPS and FEV-RIPS. During 2024–2025, the platform supported econometric expenditure analyses, multi-source information contrasts, responses to Constitutional Court judicial mandates, and publication of interactive dashboards publicly available on the ADRES institutional site. Integration of conversational AI agents (Genie, Copilot) enables analytical access for users without SQL knowledge, expanding the platform’s institutional reach.

**Conclusions:** ADRES built in one year an analytical infrastructure that provides, to our knowledge, the first published documentation of the systemic technical challenges of integrating heterogeneous data sources in a middle-income social security health system. The case demonstrates that centralizing health system information at national scale is technically feasible under the institutional constraints of a public entity — but it requires solving a set of cross-source data standardization problems that the literature on health information system implementation in middle-income countries does not document with quantitative precision. The derived lessons are transferable to health system resource administrators in Latin America facing equivalent challenges of heterogeneous information integration.

## 1. Introduction

Public health resource administrators in middle-income countries occupy a paradoxical position with respect to data. They concentrate the financial flows of systems covering tens of millions of people, yet have historically lacked the infrastructure to process that information at scale, with governance, and in operational time. The practical consequence is well documented: dependence on third parties to analyze the system they finance, inability to detect billing anomalies in operational time, and policy decisions made on aggregates constructed with months of lag [1, 2].

Colombia is no exception. The General Social Security Health System (SGSSS) covers more than 52 million people through a mixed insurance scheme in which Health Promotion Entities (EPS) receive a Per-Capita Payment Unit (UPC) from ADRES for each enrollee. ADRES also executes direct payments to providers and IPS, making it the financial hub of the system. Its theoretical access to data is broad: from enrollment through service delivery, billing, and payment. Until 2023, however, that access did not translate into operational analytical capacity. Processing billing data depended on manual uploads, fragmented schemas, and computational capacity that did not scale to the real volume of the system [3].

Resolution 2275 of 2023 from the Ministry of Health and Social Protection (MSPS) turned this capacity deficit into an urgent problem. The regulation established the gradual mandatory reporting of the Individual Health Services Delivery Record (RIPS) as supporting documentation for the Electronic Health Invoice (FEV), validated through the Single Validation Mechanism (MUV) in a new data scheme called FEV-RIPS. Implementation was organized in three groups of providers by complexity high, medium, and low — with full operation planned from June 2025 for the more than 55,000 active providers in the country [4, 5]. Decree 228 of 2025 regulated the Integrated Financial and Healthcare Information System (SIIFA) and explicitly included ADRES as an agent within its scope [6].

Building this analytical capacity faced technical challenges across multiple dimensions. ADRES had to integrate heterogeneous sources with diverse schemas, formats, and quality levels: relational databases (BDUA, direct payment records), flat files (classic RIPS), and XML documents in gradual implementation (FEV-RIPS). It also had to resolve standardization problems across sources that were not technically documented: incompatible municipality codes between enrollment and service delivery databases, extreme dates used as null value representations, evolving procedure catalogs without explicit versioning. The FEV-RIPS case illustrates the complexity: it is not an incremental extension of classic RIPS but a source with structural changes in the data schema requiring specific parsing capabilities and transformations. For ADRES, receiving and integrating these flows at national scale required solving, in parallel, two distinct problems: building the computational infrastructure that would scale to the system’s volume, and building the cleaning, normalization, and integration layers that would convert noisy, heterogeneous data — with quality problems detectable only during analytical processing into coherent, analytically usable information.

We document how ADRES resolved that problem between 2024 and 2025. What we describe is not the adoption of a pre-existing information system but the construction from scratch of an institutional analytical platform capable of processing and integrating the full universe of Colombian health system information. The scientific contribution rests on three dimensions: (i) the characterization of real heterogeneous data integration and data quality problems faced by a public resource administrator centralizing information from multiple sources in an analytical data lake — a type of problem the existing literature addresses from the provider’s perspective or from general governance frameworks, but does not document with operational technical specificity from the perspective of a national-scale administrator in a middle-income country [8, 9]; (ii) the description of technical architecture and governance decisions that allowed scaling processing to national level in one year; and (iii) the operational performance parameters that serve as a reference for administrators in the region facing similar mandates for building institutional analytical capacity [10].

## 2. Methods

### 2.1 Study design

This is a technical-descriptive report based on operational metrics from the ADRES institutional analytical platform. Its contribution does not rest on an experimental design but on the rigorous, reproducible documentation of an infrastructure implementation at a public health fund administrator, with quantitative analysis of its performance parameters and the data integration and quality problems encountered. This type of report has precedent in health informatics literature as a mechanism for transferring knowledge about implementations that would otherwise remain as unpublished institutional tacit knowledge [11].

### 2.2. Data sources and analysis period

The reported metrics come from the ADRES production environment on the Azure/Databricks platform during January–November 2025. Specific sources include: (i) pipeline execution logs in Azure Data Factory (ADF); (ii) Unity Catalog records, which document managed catalogs, tables, and records; (iii) execution duration and rows-read metrics by functional group, extracted from the platform monitoring system; (iv) records of cluster sizing deployed during the period; and (v) processing and transformation logs from multiple health system sources.

### 2.3. Architecture description

For standardization of billing data, ADRES initiated adoption of the Tuva Health Common Data Model, an open claims standardization platform designed to produce auditable, reproducible analytical tables. The implementation adapted the model to the Colombian FEV-RIPS standard, producing the highest-volume catalog in the platform (41.2% of total records). TUVA tables are views over native catalogs and do not generate double-counting in the reported total. Full adaptation of the model to the SGSSS context is under active development and constitutes an ongoing methodological work line.

The infrastructure is described through its functional components organized in layers: connectivity and transfer, automated ingestion, data lake processing, governance, and visualization/consumption. We document the data flow from on-premise and external institutional sources through to analytical outputs. For each layer we describe the implemented technical capabilities, design decisions, and the institutional constraints that determined them.

#### Connectivity and transfer capabilities

Managing massive data volumes requires mechanisms that ensure reliable connectivity and transfer. We describe: (i) file transfer and exchange mechanisms with the Ministry of Health and Social Protection; (ii) secure file upload and exchange between ADRES internal networks and the public cloud via private VPN connection protocols; (iii) capacity to process data from any source and file type without requiring prior transformation by the originating sources.

#### Data lake concept and automation

The architecture implements the data lake concept: a centralized repository that ingests data in its native format and progressively transforms it toward higher analytical quality states through medallion layers (Bronze, Silver, Gold). Ingestion and transformation pipelines are automated to the extent possible to ensure data is available in the data lake with the periodicity defined and required by each source, eliminating dependence on manual uploads. Implementing this concept is critical for ADRES’s capacity to operate with analytical autonomy over data whose format and structure it does not control.

### 2.4. Characterization of data integration and quality

Integrating heterogeneous sources into an analytical data lake surfaces quality and standardization problems detectable during processing, not during initial data receipt. We characterize three dimensions.

#### First dimension: format heterogeneities

Different sources deliver data in distinct formats (relational databases, flat files, XML documents). In the specific case of FEV-RIPS (electronic invoices delivered by MinSalud after MUV validation), processing through the ADRES-developed XML parser faces structural heterogeneities: variable XML node depth across providers of different complexity, optional fields inconsistently populated, and procedure catalog evolution without explicit versioning in the documents.

#### Second dimension: lack of standardization across sources

Integrating multiple system databases exposes coding incompatibilities: municipality codes incompatible between BDUA (enrollment) and service delivery records; extreme dates (1900-01-01, 9999-12-31) used as null value representations that generate errors in temporal aggregation operations; absence of complete technical documentation on data structures, requiring reverse engineering. We also document standardization efforts via interoperability protocols such as HL7 for BDUA consumption.

#### Third dimension: structural incompatibility between source versions

Schema differences between classic RIPS (pre-2024 historical source) and FEV-RIPS prevent direct comparability and require non-trivial transformations to build coherent longitudinal series.

For each type of problem we report: technical description of the problem, transformation implemented to resolve it, and frequency or volume of affected records.

### 2.5. Operational performance indicators

We analyze four dimensions: volume (number of records and tables managed by source), speed (processing time for massive batches), capacity (maximum computational resources deployed), and concurrency (daily query distribution by functional group). These dimensions correspond to standard evaluation metrics for large-scale data platforms [12].

### 2.6. Ethical considerations and data availability

The analyses described in section 3.7 were carried out by staff from ADRES’s Directorate of Innovation, Analytics, and Knowledge Management in the exercise of verification, control, and technical support functions assigned by the follow-up Orders to Constitutional Court Ruling T-760 of 2008 (among others, Orders 006, 875, 981, and 1245 of 2024; and 007 and 504 of 2025), and in compliance with Resolutions 370 and 1324 of 2025 from the Ministry of Health and Social Protection, which establish ADRES’s institutional participation in the Technical Working Group for UPC review. Access to individual financial and billing information was carried out under explicit legal mandate, in secure internal environments with role-based access controls. All reported results are exclusively aggregated. This technical report does not constitute research with human subjects and did not require external ethics committee approval.

## 3. Results

ADRES’s analytical infrastructure was built on Microsoft Azure as the cloud provider, with Databricks as the central distributed processing engine. The architecture implements a multi-source data lake with a three-layer medallion pattern (Bronze, Silver, Gold) [27]. This design responds to a structural constraint: ADRES does not control the formats in which external sources deliver data, so integration capacity must be built internally without requiring prior standardization.

Bronze stores data in native format — XML, relational tables, flat files — and accepts residual heterogeneities and errors. Silver applies cleaning and homologation transformations: conversion to standard schemas, elimination of extreme dates used as nulls, validation of codes against reference catalogs, and correspondence tables between sources with incompatible coding. Gold contains integrated, denormalized data ready for analytics, combining information from multiple sources crossed via validated keys. The separation between layers ensures that problems in Bronze do not contaminate the analytical layers.

On-premise sources connect to the cloud environment via VPN Gateway with Self-Hosted Integration Runtime (HTTPS, port 443), keeping institutional databases within the ADRES internal network. For exchange with the Ministry of Health, ADRES established direct connectivity via Microsoft Fabric OneLake data shortcuts, without intermediate transfers.

Azure Data Factory and Databricks orchestrate fully automated pipelines by source type, with frequencies defined by the data (daily for BDUA, monthly for UPC settlements). Each execution generates logs with full traceability. Automation eliminated the manual uploads that prior to 2024 generated multi-day lags and referential integrity errors.

Databricks manages transformations through standard connectors for relational data and a specialized Python/PySpark module for FEV-RIPS XML parsing, which extracts structured information from electronic invoices while resolving inter-provider heterogeneities: variable node depth, inconsistent optional fields, and catalog evolution without explicit versioning. Microsoft Fabric is integrated for machine learning models and predictive analytics.

Unity Catalog centralizes the catalog with role-based access control (RBAC) segmented by functional group: the Audit team accesses billing tables but not individual enrollment; the UPC team accesses historical expenditure series but not patient-level detail. A Data Governance Dashboard (DGD) monitors the ecosystem in real time: outdated tables, failed pipelines, ingestion volumes by source.

The platform serves eleven functional groups through Power BI dashboards, REST APIs, shared notebooks, and automated reports. Integration of Genie (Databricks) and Microsoft Copilot (Power BI) enables natural language queries without SQL, extending analytical access to non-technical users. The Intelligence Room establishes segmented domains for sharing analyses with external system institutions. Public dashboards with downloadable micro-data (Direct UPC Payment, Maximum Budgets, services to deceased persons) extend access to researchers and the public.

### 3.2. Technical challenges in heterogeneous data integration

Building a data lake that integrates multiple Colombian health system sources in diverse formats exposed technical challenges that consumed development time not anticipated in the initial project estimate. These challenges fall into three categories: format heterogeneities within individual sources, lack of standardization across system sources, and structural incompatibility between historical and current versions of the same sources.

#### 3.2.1. Processing unstructured data: the case of Electronic Health Billing (FEV-RIPS)

In the billing evaluation that ADRES conducts in support of the Ministry of Health, we process XML documents from electronic invoices that have passed technical validation through the Single Validation Mechanism (MUV). The ADRES-developed parser extracts structured information from these documents and transforms it into relational tables within the data lake. This processing faces heterogeneities derived from differences in standard implementation across providers with different technical capacities.

##### (i)Variable XML node depth

High-complexity providers (university hospitals, specialized clinics) generate documents with deep nested node structures to record complex procedures with multiple components. Low-complexity providers (outpatient clinics, primary care centers) generate flat-structure documents where each service is a first-level node. The parser must extract information from both structures without data loss or assignment errors between parent-child nodes.

##### (ii)Optional fields inconsistently populated

The FEV-RIPS schema defines optional fields (secondary diagnosis, related procedure, clinical observations) that some providers populate systematically and others leave empty or use as unstructured free text. This generates tables with a high proportion of null values requiring specific treatment to prevent errors in aggregation and filtering operations.

##### (iii)Procedure catalog evolution

Procedure coding uses the CUPS catalog (Unique Classification of Health Procedures), updated periodically by MinSalud. During 2024–2025, documents with codes from different CUPS versions coexisted without an explicit field identifying the version used. We developed institutional equivalence tables that homologate codes across versions to enable coherent longitudinal series.

#### 3.2.2. Lack of standardization across system sources

Integrating multiple system databases exposed coding incompatibilities and quality problems detectable only during cross-source analytical processing.

##### (i)Incompatible municipality codes

Municipality codes used in BDUA (enrollment) do not consistently match codes used in service delivery records (RIPS, FEV-RIPS). Some municipalities have different codes in each database; others appear in one but not the other. This prevents direct joins for territorial expenditure analysis. ADRES developed an institutionally maintained equivalence table mapping codes across sources, updated when new inconsistencies are detected during processing.

##### (ii)Extreme dates as null representations

Multiple system databases use extreme dates (1900-01-01, 9999-12-31, 1899-12-30) as representations of missing values in date-type fields. This generates errors in temporal aggregation operations (days-between-events calculations, monthly series construction) and requires cleaning routines that detect and replace these dates with explicit nulls before analytical processing. The absence of technical documentation about this widespread practice extended debugging time for failed pipelines.

##### (iii)Absence of complete technical documentation

Structured databases delivered by external sources lack exhaustive technical documentation on field structure, valid value catalogs, and implemented business rules. This requires reverse engineering: exploratory analysis of value distributions, primary and foreign key identification via uniqueness and reference analysis, and validation rule inference through pattern detection. This process consumes specialized technical development time and creates risk of incorrect interpretations of field meaning.

##### (iv)Standardization and interoperability efforts

We document institutional efforts to improve source interoperability through standard protocols such as HL7, particularly for structured BDUA consumption. These efforts are complementary to the transformations implemented in the data lake but are under development and do not retroactively resolve historical data incompatibilities.

**Table 1.** Data integration problems identified during ADRES analytical data lake processing, 2024–2025.

| Integration problem identified | Affected sources | Implemented transformation |
| --- | --- | --- |
| Incompatible municipality codes | BDUA; classic RIPS; FEV-RIPS | Institutional equivalence table maintained by ADRES, updated upon detection of new inconsistencies during processing |
| Extreme dates as null representations (1900-01-01; 9999-12-31; 1899-12-30) | Multiple system sources | Detection and replacement routines with explicit NULL before analytical processing in Silver layer |
| Optional fields empty or with free text (secondary diagnosis; clinical observations) | FEV-RIPS | Null treatment in transformation pipelines; normalization in Silver layer |
| Absence of complete technical documentation (fields; catalogs; business rules) | External sources (MinSalud; EPS) | Reverse engineering: exploratory distribution analysis; key identification via uniqueness analysis; validation rule inference |
| Schema incompatibility between classic RIPS and FEV-RIPS (absent financial fields; ICD-9 to ICD-10 migration) | Classic RIPS; FEV-RIPS | ICD-9/ICD-10 correspondence tables; differentiated analysis logic by source type by period |
| CUPS catalog evolution without explicit versioning in documents | FEV-RIPS | Equivalence tables across CUPS versions; homologation in Silver layer |
| Variable XML node depth across providers of different complexity | FEV-RIPS | Python/PySpark parser with recursive node extraction; parent-child assignment validation |

*Note: Transformations are applied in the Silver layer of the medallion pattern. Source: development and pipeline monitoring records, ADRES, 2024–2025.*

#### 3.2.3 Structural incompatibility between source versions

Classic RIPS (the historical service delivery information source through 2023) and FEV-RIPS are not directly comparable. The differences are not only in format (flat files versus XML) but in granularity, coverage, and data schema.

##### (i)Financial fields absent in historical version

FEV-RIPS incorporates detailed financial information billed value, authorized value, applied discounts, copayments, and deductibles — that classic RIPS did not contain. This prevents longitudinal analyses of billing financial components that require comparing pre- and post-2024 periods.

##### (ii)Diagnosis coding catalog migration

The diagnosis coding scheme migrated from ICD-9 to ICD-10 between versions, without an explicit field identifying the version used in each record. Some procedure coding also changed. We developed ICD-9 ↔ ICD-10 correspondence tables to homologate diagnoses in historical series.

##### (iii)Longitudinal series construction strategies

ADRES implemented analysis-specific logic: aggregated series (consultation volume, hospital discharges) combine both sources with non-duplication validation via composite keys; individual-detail analyses (service traceability by patient) use only post-2024 FEV-RIPS to ensure schema consistency; methodological change analyses explicitly compare both sources in overlapping periods to quantify the impact of structural differences.

Technical validation of these transformations consumed substantial development time. Documentation of the structural incompatibility problem between versions of health registry standards is not present with sufficient technical detail in the literature on information system implementation in middle-income countries, which required designing solutions without established methodological references.

### 3.3. Volume and variety of data managed

As of November 2025, the platform manages 21 production data catalogs, 1,183 tables, and approximately 110,645 million stored records. Cumulative processing output production exceeds 1 trillion records. The highest-volume catalogs correspond to electronic billing, RIPS, FEV-RIPS, BDUA, and sufficiency, reflecting the densest information flows of the SGSSS [see Figure 1].

**Figure 1.**
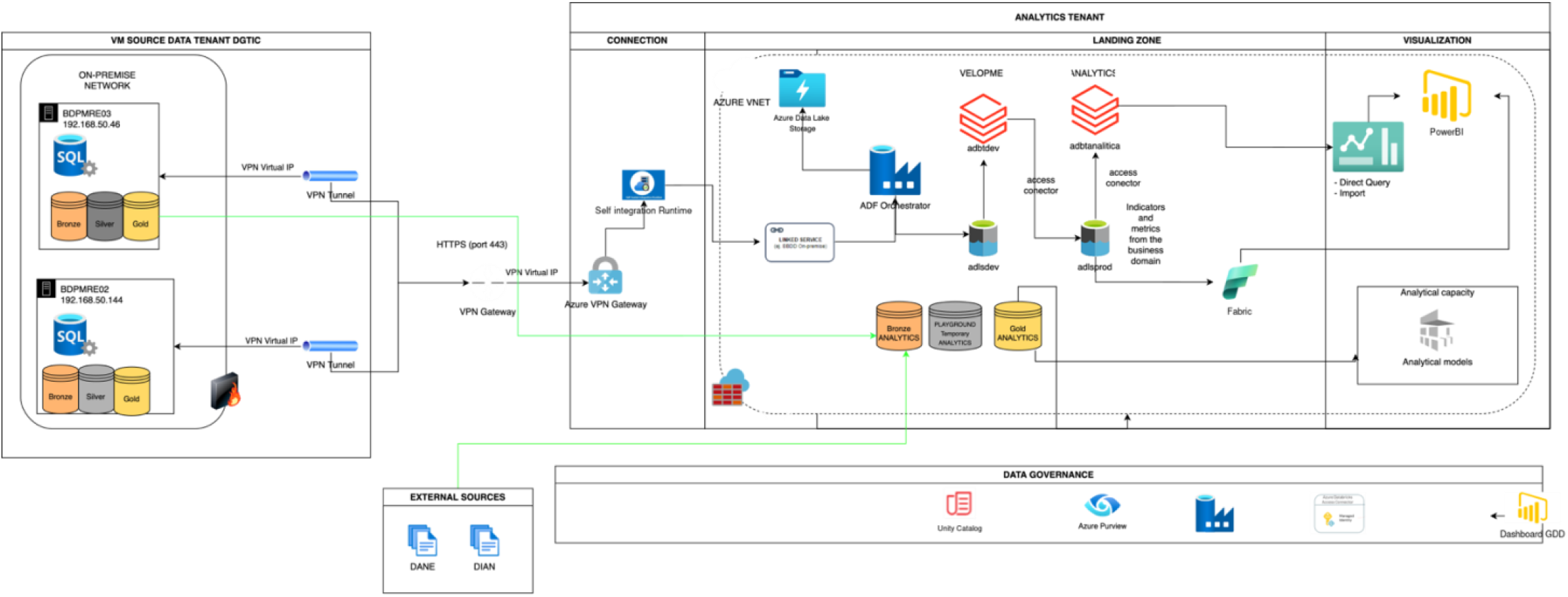
Data engineering implementation architecture — ADRES, 2025. Full technology stac diagram: on-premise sources, VPN/Azure connectivity, orchestration (ADF), distributed processing (Bronze/Silver/Gold in Databricks), governance (Unity Catalog, Azure Purview, GDD), and visualization (Power BI, Fabric). Source: ADRES, 2025.

**Table 2.** Production data catalog inventory. ADRES analytical platform, November 2025.

| Catalog | Tables | Records (millions) | % of total | System source |
| --- | --- | --- | --- | --- |
| TUVA | 183 | 45,538.3 | 41.2% | Tuva Health Common Data Model — standardized views over native catalogs (FEV-RIPS, RIPS, electronic billing) |
| UPC Sufficiency | 45 | 22,231.5 | 20.1% | UPC sufficiency calculation and maximum budgets |
| BDUA | 20 | 16,913.1 | 15.3% | Unified Enrollee Database |
| Classic RIPS | 117 | 6,136.0 | 5.5% | Individual service delivery record pre-2024 |
| Electronic billing | 167 | 5,569.7 | 5.0% | Electronic billing UPC technical working groups and verification |
| FEV-RIPS | 54 | 4,964.8 | 4.5% | Electronic Health Invoice with embedded RIPS (Res. 2275/2023) |
| MIPRES | 212 | 4,379.1 | 4.0% | Prescription of medications and technologies outside the PBS |
| UPC | 19 | 3,952.6 | 3.6% | Per-Capita Payment Unit recognition and historical series |
| Claims | 18 | 507.6 | 0.5% | Claims and billing disputes between EPS and IPS |
| PVS | 41 | 297.5 | 0.3% | Voluntary Health Plans |
| Master data | 52 | 80.4 | 0.1% | Institutional reference catalogs |
| Direct payment | 8 | 29.3 | 0.0% | Direct payment to IPS and payment series |
| BDEX | 4 | 14.3 | 0.0% | Exclusions database |
| SIVIGILA | 1 | 10.2 | 0.0% | Public Health Surveillance System |
| SIA | 80 | 10.0 | 0.0% | Outpatient Information System |
| REPS | 12 | 5.1 | 0.0% | Special Registry of Healthcare Service Providers |
| DGRF | 4 | 4.1 | 0.0% | General Directorate of Financial Regulation |
| Anomalies | 82 | 1.3 | 0.0% | Anomaly detection models and intelligent auditing |
| System | 47 | 152k | 0.0% | Platform metadata and data governance |
| RETHUS | 1 | 45k | 0.0% | Health Human Resources Registry |
| DataGov | 16 | 18 | 0.0% | Institutional data governance |
| Total production | 1,183 | 110.6B | 100.0% | 21 active catalogs |

*Note: Development and testing catalogs excluded. Source: Unity Catalog, ADRES, 2025.*

The variety of sources feeding the platform is technically relevant beyond volume. Format heterogeneity required developing specific connectors for each source type and normalization layers that homologate them in the institutional data model before they can be joined for analysis.

### 3.4 Processing speed

The platform can process 220 million electronic invoices in three hours under normal operating conditions [see Figure 2]. It executes thousands of distributed queries across eleven functional groups, with monthly peaks of 78,028 queries in October 2025, coinciding with system settlement and audit cycles. The Databricks distributed processing architecture allows horizontal scaling to absorb these peaks without structural redesign.

**Figure 2.**
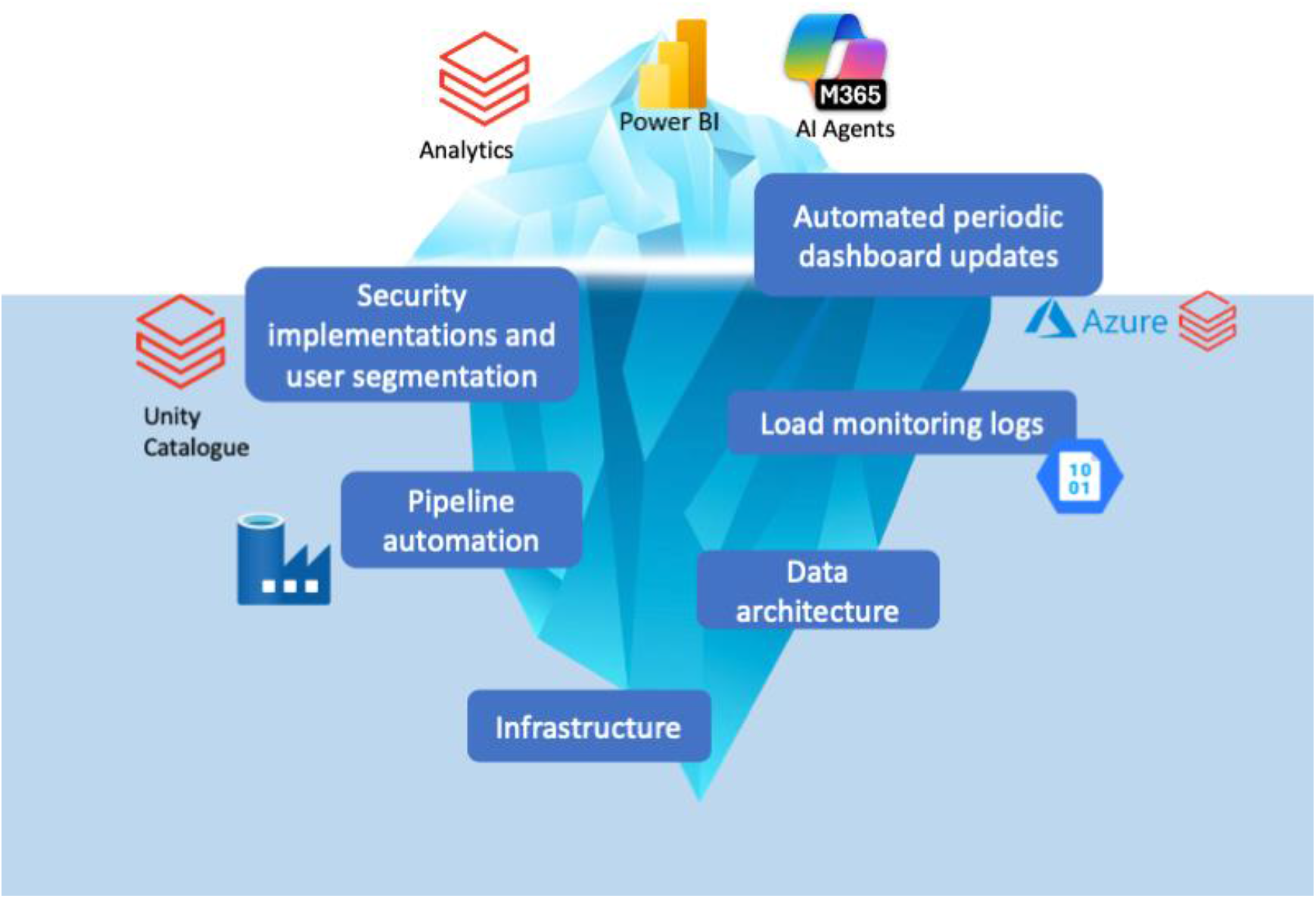
Components of institutional DataOps — ADRES, 2025. Iceberg representation of visible components (analytics, Power BI, AI agents) versus underlying infrastructure (data architecture, pipeline automation, monitoring, security, Unity Catalog). Source: ADRES, 2025.

**Table 3.** Query activity on the ADRES analytical platform, January–December 2025.

| Month | Queries executed | % of total | Operational context |
| --- | --- | --- | --- |
| January 2025 | 73 | 0.0% | Operations start |
| February 2025 | 9,458 | 2.6% |  |
| March 2025 | 43,912 | 12.0% | Intensive analytical cycle |
| April 2025 | 37,470 | 10.3% |  |
| May 2025 | 25,554 | 7.0% |  |
| June 2025 | 17,461 | 4.8% |  |
| July 2025 | 28,463 | 7.8% |  |
| August 2025 | 19,042 | 5.2% |  |
| September 2025 | 51,098 | 14.0% | Audit cycle |
| October 2025 | 78,028 | 21.4% | Annual peak: settlement and audit |
| November 2025 | 16,241 | 4.5% |  |
| December 2025 | 37,845 | 10.4% | Year-end close |
| Total 2025 | 364,645 | 100.0% | 11 institutional functional groups |

*Note: Queries executed on Databricks SQL Warehouse environment. September and October coincide with UPC settlement and billing audit cycles. Source: Databricks monitoring system, ADRES, 2025.*

The capacity to process the full universe of records has methodological consequences for audit functions. Anomaly detection models operating on samples risk not detecting patterns visible only at scale. The ADRES platform eliminates that constraint for SGSSS billing analysis.

### 3.5 Computational capacity

The infrastructure deploys clusters of up to 32 TB RAM and 4,096 vCPU, a scale that enables executing queries on 100 billion records in ten seconds [see Figure 3]. Sizing is elastic: clusters scale according to demand and reduce outside peak periods, meaning operational cost is a function of actual use rather than permanently reserved maximum capacity.

**Figure 3.**
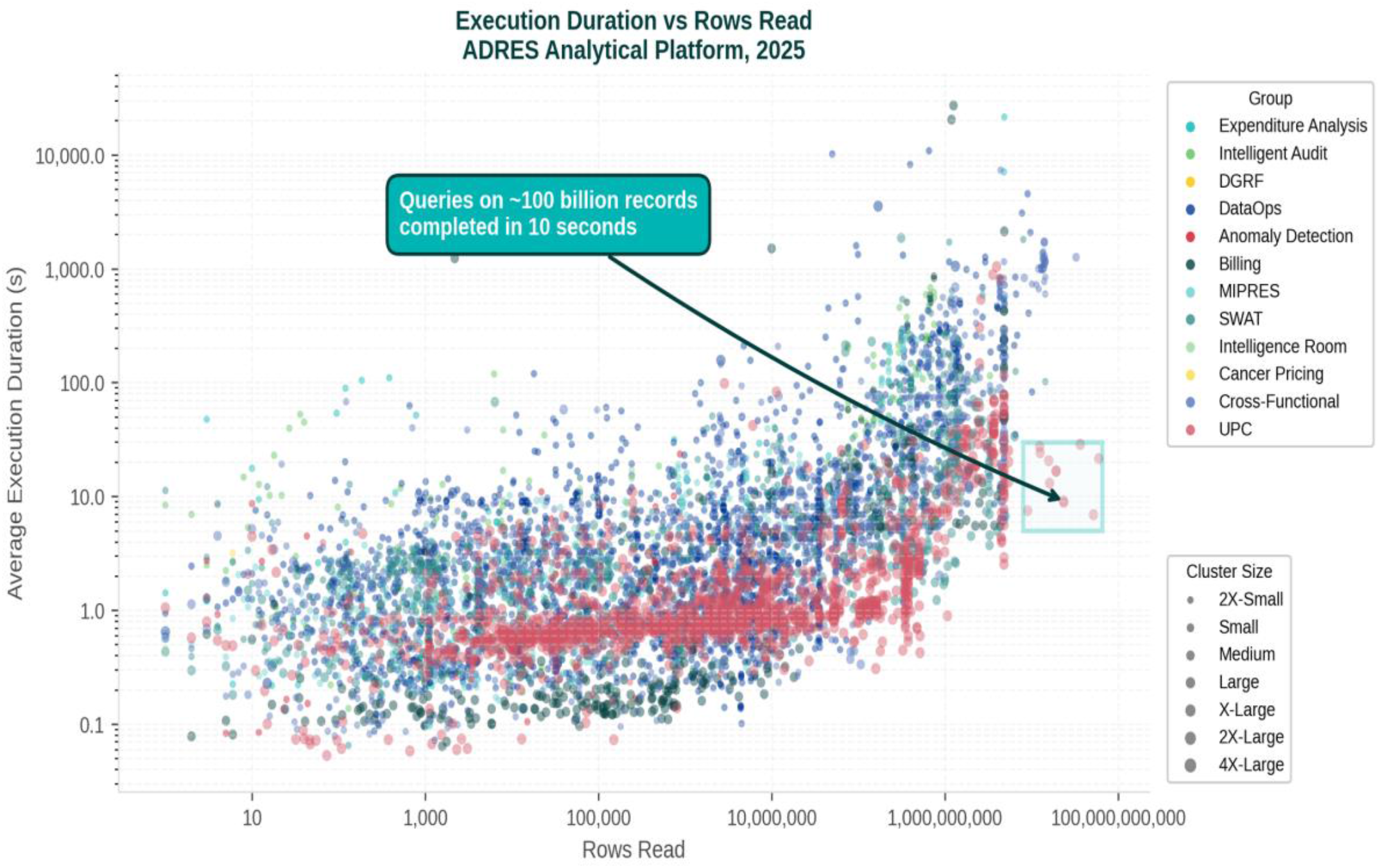
Volume and capacity of the ADRES analytical platform, January–November 2025. Left: number of tables and records by catalog (21 catalogs, 1,183 tables, 110,645 million stored records; production >1 trillion records). Right: execution duration versus rows read by functional group; queries on ∼100 billion records completed in 10 seconds. Source: ADRES, 2025.

**Figure 4.**
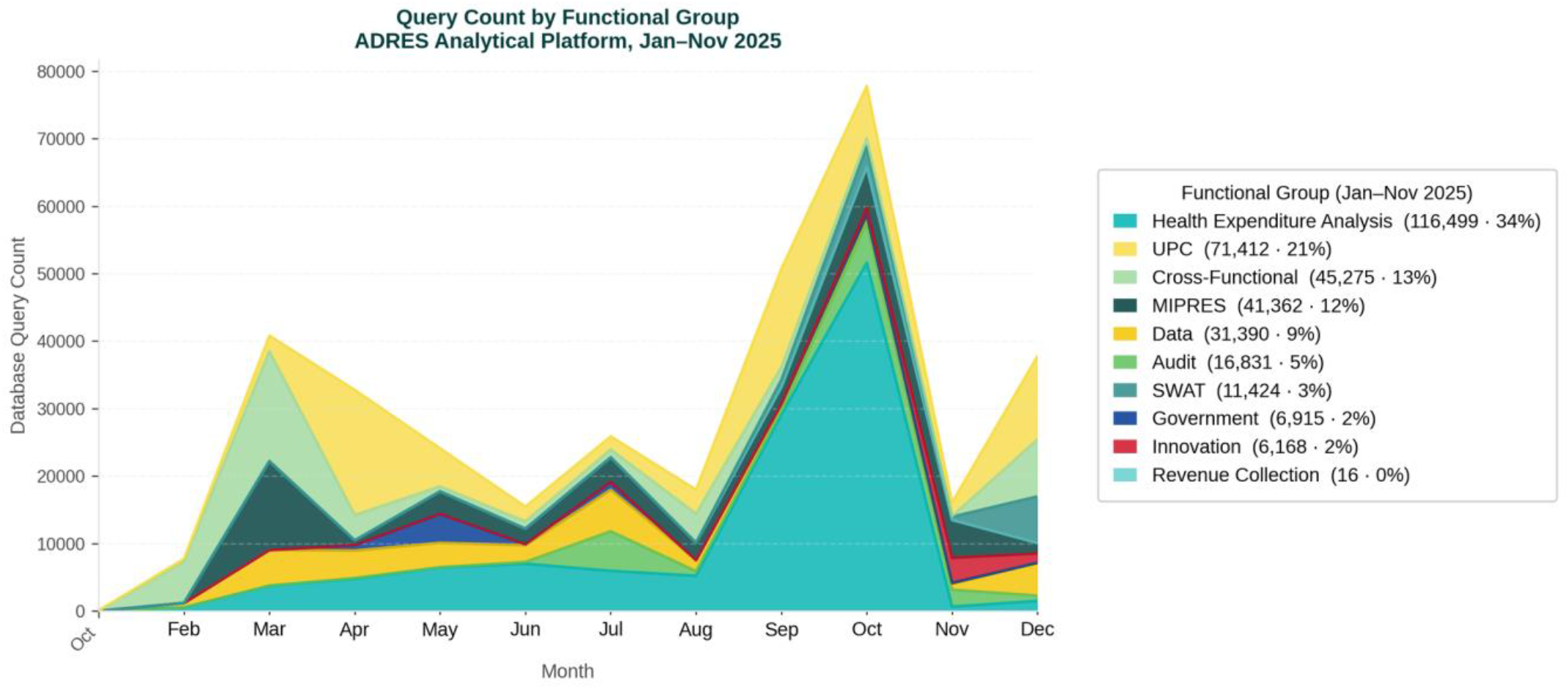
Speed and concurrency — ADRES, January–November 2025. Left: daily query count by functional group, with peak of 78,028 monthly queries (October 2025). Right: processing indicators: 220 million invoices in 3 hours; clusters of up to 32 TB RAM and 4,096 vCPU. Source: ADRES, 2025.

### 3.6 Data governance: a challenge under development

Operating a data lake that integrates clinical and financial information on millions of people requires a data governance model that guarantees privacy, security, and accuracy, in accordance with the requirements established in Article 2.12.1 of Decree 780 of 2016 as updated by Decree 228 of 2025 [6].

The platform implements Unity Catalog as the basis for access control, with functional group segmentation restricting which users can query which data lake tables. An institutionally developed Data Governance Dashboard (DGD) monitors the ecosystem state: outdated tables, failed pipelines, ingestion volumes by source. These components establish basic operational access and traceability controls.

However, building a formal data governance model that includes fully documented metadata, automatic transformation lineage, sensitive information classification, and differentiated retention policies by data type is a challenge under active development. The nature of the Colombian health system — with heterogeneous sources developed independently over decades — requires that governance be built progressively over an already-operational infrastructure, rather than designed prior to implementation. This formalization process is ongoing and constitutes a priority area of institutional development [14].

The institutional response to this challenge has a formal three-level structure. The strategic level is the Institutional Management and Performance Committee (CIGD). The tactical level is the Data Governance and Analytics Subcommittee (SGAD), responsible for quality, access, and data use policies. The operational level is the Domain Technical Working Groups that execute SGAD agreements on specific sources. The official governance maturity diagnosis places ADRES at Level 2 of 5 (Managed) on the adopted institutional scale. The pilot of the Settlement and Payments Domain (DLyG), in advanced status as of October 2025, reports 100% of roles defined, 100% of flows documented, 90% of master data cataloged, and 80% of glossary completed. Priority domains for 2025– 2026 are BDUA, Direct Payment, and the Intelligence Room. Pending components include formal data quality auditing, extension of the model to other directorates, and progressive maturity toward Level 3.

### 3.7 Analytics

The analyses reported in section 3.7 were executed on data from the curation layer (Gold) of the medallion pipeline for BDUA, MIPRES, sufficiency, direct payment, and master data sources. Analyses integrating RIPS (Silver layer) used the institutional municipality code homologation table documented in section 3.2. Formal organizational governance was under active development during this period, which constitutes a recognized limitation of the infrastructure in its current stage.

The analytical products described below were developed by the ADRES Analytics team using the described infrastructure. Their mention constitutes evidence of the platform’s operational scope, not a methodological validation of the individual analyses.

In compliance with Constitutional Court Orders 007 and 504 of 2025, ADRES produced a technical report identifying medical services billed after the enrollee’s date of death in the period 2018–2023, corresponding to 471,480 individuals for a total of COP 2.3 trillion. The findings were publicly communicated and formally reported to the Attorney General’s Office.

The platform described in the preceding sections made it possible during 2024 and 2025 to execute a set of health system expenditure analyses that were previously not executable — not for lack of research questions, but because the sources needed to answer them were not centralized in an environment where they could be crossed at full scale. That these exercises could be executed over the course of a year, several with publication of interactive micro-data, is a direct result of the infrastructure and the integration of artificial intelligence tools in analytical processes.

The developed products are organized in three lines. The first line comprises technical health expenditure analytics reports requiring multi-source joins and full-universe record processing: analysis of UPC-financed health expenditure under the efficiency principle of Law 100 (2018–2023); econometric determinants of UPC expenditure via regression model (fiscal year 2023); analysis of UPC-related information sources and average cost per enrollee and its historical behavior; effect of voluntary health plan enrollment on UPC-financed service costs (2018–2023); insurer market concentration via Herfindahl-Hirschman Index (2009–2024), identifying changes in the sector’s competitive structure and risks associated with loss of competition. These reports share a technical characteristic: none can be executed on a single source; all require distributed joins across tables from different origins, cross-source key validation, and capacity to process the full universe of records without sampling.

The second line comprises compliance reports for judicial mandates and information contrasts [24, 25]. During 2025, the Constitutional Court issued Orders 007, 089, and 504, requiring analysis of EPS-issued authorizations and contrast of medication records against other official databases. The contrast between service delivery records and the deceased enrollment status in BDUA (2018–2023) identified services billed after the enrollee’s date of death, generating technical inputs to improve information reporting quality and the proper allocation of public resources [23]. The Financial Working Group final report contrasted information between EPS financial reports and other official sources (2019–2023), evaluating its impact on UPC resource utilization and contributing evidence to strengthen the quality, consistency, and traceability of system financial information. The UPC calculation contrast exercise identified differences and inconsistencies between values reported by EPS and ADRES institutional databases, aimed at improving the precision and traceability of the calculation [26].

The third line comprises interactive dashboards publicly available on the ADRES institutional portal. The Direct UPC Payment dashboard presents the 2020–2026 series disaggregated by regime, EPS, provider, and department; total direct payment exceeded COP 68.7 trillion in 2025. The Maximum Budget Monitoring dashboard reports for 2024: COP 4.25 trillion in supply value, 748,368 patients, 4.78 million records, with analysis by technology, population profile, EPS, IPS, prescriber, diagnosis, and alert generation. The services-to-deceased-persons contrast dashboard enables queries by activity, technology, and days between care and date of death, disaggregated by EPS and by individual. Integration of conversational AI agents allows users without SQL or Python knowledge to formulate natural language questions about the data and receive automatic responses, democratizing analytical access beyond specialized technical teams. The Intelligence Room establishes segmented information domains with differentiated access levels, enabling secure sharing of analyses with external Colombian health system institutions that currently use it in their decision-making processes.

## 4. Discussion

We document the construction of a centralized data infrastructure for health expenditure analysis in a middle-to-high-income national system. The results demonstrate that resolving analytical fragmentation through cloud-native architectures with distributed processing is technically feasible, even when information sources are heterogeneous, coding standards are inconsistent, and technical documentation is incomplete or absent. The scale of centralization — 110 billion records across 21 production catalogs — and the capacity to execute cross-source queries on the full national billing universe establish an operational reference point for other public administrators facing similar challenges.

### 4.1 Integration challenges: beyond format problems

The literature on electronic billing standard implementation systematically describes infrastructure, interoperability, and adoption barriers [8, 9, 15, 21]. What that corpus documents with less precision is the operational dimension: what inconsistencies persist after formal validations have been passed, and what transformations are required for data to be analytically usable.

section 3.2 documents three problem categories. Format heterogeneities within individual sources are the type the literature records most frequently [7, 8]. Lack of standardization across sources and structural incompatibility between historical versions concentrated the largest share of technical effort in this case and receive less attention in the international literature. A representative example: municipality codes in BDUA and in service delivery records are each valid within their own schemas but incompatible with each other, and no official correspondence table exists — ADRES builds and maintains one institutionally. The absence of technical documentation on catalogs and business rules forces reverse engineering through distribution and uniqueness analysis, with risk of incorrect interpretations of field meaning.

The incompatibility between classic RIPS and FEV-RIPS is not a format migration problem. It is partial semantic incompatibility affecting the capacity to build longitudinal series and forcing design decisions about which version of the data to use for each type of analysis. The development time for the transformation routines was not anticipated in the initial estimate. That is probably the most underestimated component in equivalent projects in the region.

### 4.2 The administrator’s responsibility as sovereign processor

The platform enabled analyses that were not executable with isolated sources. One documented example is the identification of medical services billed after the enrollee’s date of death in the period 2018–2023, corresponding to COP 2.3 trillion, published in the ADRES Analytics Directorate’s contrasting report [23]. This type of analysis, executed by the analytics team under ADRES’s legal mandate, illustrates the link between integrated analytical infrastructure and effective public health expenditure control.

Decree 228 of 2025 did not delegate a discretionary technical function to ADRES; it regulated ADRES as a SIIFA agent with specific obligations for receiving, processing, and analyzing the system’s financial and healthcare information [6]. This has implications beyond day-to-day operations. An administrator that can process the full universe of electronic billing with operational response capacity can detect anomalies that an administrator dependent on third parties or operating with months of lag cannot detect. The capacity built is, in that sense, a necessary condition for exercising the intelligent audit function that the regulatory framework assigns.

The financial stewardship literature holds that an administrator’s capacity to see the full system is determinative for expenditure control and equitable resource distribution [16, 20]. The infrastructure described in this paper is the technical component that enables that capacity in the Colombian case. section 3.7 documents concrete analytical products executed during 2024 and 2025 that required this infrastructure: analysis of services billed after enrollee death, contrasting EPS-reported values against ADRES institutional databases, responses to Constitutional Court mandates within judicial deadlines. These capacities did not exist before 2024; what changed was not the research question but the technical capacity to answer it at full scale with verifiable traceability.

### 4.3 Transferability hypotheses for public health fund administrators in Latin America

The hypotheses formulated below derive from the experience documented in this paper and represent verifiable propositions for future comparative research. They do not constitute confirmed generalizations or ADRES institutional positions on regional health policy.

Platform deployment was transparent to providers in terms of additional transmission requirements. However, centralization generated, for the first time at national scale, quantitative evidence of the magnitude of data quality problems in data providers were already producing. This creates the technical conditions for grounding data quality mandates with endogenous evidence, and for designing technical support programs for IPS — particularly lower-complexity providers — as a condition for improvement to be systemic rather than merely normative.

The ADRES experience is not directly transferable as a technological model, since each entity operates under distinct budgetary, regulatory, and inherited infrastructure constraints — but it does permit formulating hypotheses about design decisions whose relevance may be independent of specific context.

#### H1. Separating the raw ingestion layer from analytical layers as a noise management mechanism

The medallion pattern (Bronze/Silver/Gold) allows data with format errors or inconsistencies to reach the Bronze layer without contaminating the Silver and Gold layers, which feed analysis models and decision dashboards. This architecture enables processing to continue even when input data has quality problems, without those problems propagating to final analytical products.

#### H2. Data governance as a progressive development component, not an initial blocker

Implementing Unity Catalog and the Data Governance Dashboard establishes basic access and traceability controls from the start, but building the complete formal governance model is a process that develops in parallel with infrastructure operation. The alternative — waiting for governance to be fully formalized before putting infrastructure into production — would have delayed institutional analytical capacity by years.

#### H3. Prioritizing multi-source processing capacity over individual source perfection

Cross-source standardization problems (municipality codes, extreme dates as nulls, absent documentation) are not resolved by building analytical infrastructure; they are resolved through inter-institutional coordination and standard adoption — processes operating on longer timescales than technical projects. The decision to build institutional integration capacity allows operational progress while standardization efforts mature.

#### H4. Integration of conversational AI tools (Genie, Copilot) enables analytical access for non-technical users

Using these tools to guide non-technical users in SQL expands the institutional reach of the platform, but this democratization is only possible because an underlying infrastructure guarantees data quality, security, and governance. AI does not replace the infrastructure; it amplifies its institutional reach.

### 4.4 Limitations

We document the implementation of a specific infrastructure in a specific institutional context; the transferability of the technical solution to other public administrators will depend on factors including budget availability for cloud services, internal technical capacity or external contracting possibilities, regulatory framework on health data privacy and security, and characteristics of the data source ecosystem in each jurisdiction. The reported metrics correspond to the first full year of operation (2024– 2025); medium-term performance stability remains to be documented. The analysis does not include cost-effectiveness evaluation of the infrastructure compared with alternatives (on-premise processing, hybrid solutions). This paper does not document the organizational process of platform adoption by institutional functional groups, nor the impact of data availability on public policy decision quality — these are complementary research questions requiring different methods. Comparison with other public administrators in the region is difficult due to the scarcity of published technical reports on equivalent infrastructures.

**Table 4.** Operational cost of the ADRES analytical platform by service, May 2025–April 2026 (USD).

| Month | Databricks (USD) | Storage (USD) | Defender (USD) | VMs (USD) | Total (USD) |
| --- | --- | --- | --- | --- | --- |
| May 2025 | 1,877.52 | 1,355.36 | 687.78 | 344.26 | 4,264.92 |
| June 2025 | 1,364.84 | 1,368.58 | 623.85 | 2,858.09 | 6,215.36 |
| July 2025 | 2,982.40 | 1,570.36 | 717.04 | 1,509.24 | 6,779.04 |
| August 2025 | 3,637.31 | 2,632.84 | 2,213.16 | 1,284.61 | 9,767.92 |
| September 2025 | 4,114.46 | 3,419.19 | 972.38 | 999.86 | 9,505.89 |
| October 2025 | 11,177.11 | 5,455.41 | 1,396.38 | 481.91 | 18,510.81 |
| November 2025 | 10,363.22 | 5,437.35 | 1,047.29 | 466.31 | 17,314.17 |
| December 2025 | 15,682.81 | 17,058.30 | 1,955.77 | 412.39 | 35,109.27 |
| January 2026 | 3,968.37 | 2,530.00 | 1,296.31 | 323.91 | 8,118.59 |
| February 2026 | 17,642.59 | 1,714.29 | 1,609.69 | 293.29 | 21,259.86 |
| March 2026 | 15,241.02 | 2,085.14 | 618.39 | 395.29 | 18,339.84 |
| April 2026 | 9,519.55 | 1,909.43 | 598.42 | 529.69 | 12,557.09 |
| Total | 97,571.20 | 46,536.25 | 13,736.46 | 9,898.85 | 167,742.76 |

*Note: Costs recorded in resource group grpanalitica under the Public Cloud Pricing Framework Agreement CCE-241-AMP-2021. The storage peak in December 2025 corresponds to a routine Microsoft Defender for Cloud review. Source: Azure Cost Management, ADRES, 2026.*

Total operational cost during May 2025–April 2026 was USD 167,743, distributed across Azure Databricks (USD 97,571; 58.2%), storage (USD 46,536; 27.7%), Microsoft Defender for Cloud (USD 13,736; 8.2%), and virtual machines (USD 9,899; 5.9%). Monthly expenditure ranged from USD 4,265 (May 2025) to USD 35,109 (December 2025); compute peaks (Databricks) concentrated in months of highest analytical activity (October 2025, February–March 2026), while the December peak was associated primarily with a routine Microsoft Defender for Cloud review that generated extraordinary storage charges that month. These figures do not constitute a generalizable market cost reference: acquisition of cloud services by Colombian state entities is conducted through the Public Cloud Pricing Framework Agreement of the National Public Procurement Agency — Colombia Compra Eficiente (CCE-241-AMP-2021), a demand aggregation instrument establishing maximum negotiated prices for the public sector [19]. The reported costs reflect aggregate public procurement conditions and are not directly comparable to open-market rates or entities operating under different procurement schemes. The absence of published cost benchmarks for national-scale health data platform operation in middle-income countries prevents systematic comparison; this limitation is itself an additional argument for publishing technical reports such as this one.

Three implementation process challenges merit explicit documentation as findings. First, administrative delays in information delivery by external sources affected ingestion schedules and fostered a formalization process for information exchange to ensure pipeline continuity — these pipelines being dependent on administrative processes operating outside the technical team’s control. Second, data quality was substantially lower than expected: anomalous data, absence of standardization in medication, procedure, and diagnosis names, and documentary gaps required unanticipated additional cleaning work not reflected in the initial estimate. Third, given the magnitude of the standardization deficit, the team employed artificial intelligence tools for the identification and normalization of non-standardized records. This proved an effective resource but with computational and human supervision costs that should not be underestimated in equivalent projects. These three findings do not invalidate the platform; they document the real operating conditions in a middle-income health system with decades of data produced under heterogeneous schemes.

## 5. Future Perspectives

ADRES built between 2024 and 2025 an analytical infrastructure whose operational performance constitutes, to our knowledge [22], the first published documentation of the systemic technical challenges of heterogeneous source integration in a Latin American middle-income social security system. The platform demonstrates that processing the totality of national health system information at scale, with response times compatible with operational management, is technically achievable under the institutional constraints of a public entity.

Future development of this capacity proceeds in three directions. The first is consolidation of the data governance model. Unity Catalog and the Data Governance Dashboard establish basic access and traceability controls; the next step is full model formalization: metadata documented according to institutional standards, automatic transformation lineage enabling each calculated field’s origin to be traced, sensitive information classification with differentiated retention policies by data type, and quality protocols that automatically detect degradation in input sources. This consolidation is technical but also organizational; it requires inter-institutional coordination with the entities that generate the source data.

The second direction is integration of new system information sources. The current platform processes BDUA, classic RIPS, FEV-RIPS, MIPRES, direct payments, maximum budgets, and sufficiency. Additional sources that would expand analytical capacity include: outpatient morbidity records, service supply information (REPS), occupational disability certifications, and care quality data reported by providers. Each new source replicates the integration challenges documented in section 3.2 but also expands the types of questions the resource administrator can formulate about system functioning. Interoperability standard adoption efforts such as HL7, particularly for structured BDUA consumption, are complementary to the transformations implemented in the data lake and will facilitate future source integration as they are uniformly implemented across the health system ecosystem.

The third direction is expansion of advanced analytics capabilities through machine learning techniques applied to the full universe of centralized records. The analytical products documented in section 3.7 operate on aggregations or samples of the complete data. The next capacity level is applying predictive models trained on the full universe: anomalous billing pattern detection, service demand prediction by region and population profile, high-cost care trajectory identification, and public policy scenario modeling via simulations incorporating the system’s observed historical behavior. These applications require not only computational infrastructure but also internal methodological capacity development and careful model validation before they inform policy decisions.

The least documented dimension in the literature for those facing equivalent projects in middle-income health systems is data quality and, particularly, lack of standardization across system databases. The structural incompatibility between classic RIPS and FEV-RIPS, the absence of homologated codes across sources (municipalities, providers, diagnoses), the use of extreme dates as null representations, and the absence of complete technical documentation generated a set of technical problems that consumed unanticipated development time and capacity. Documenting that problem with precision is the primary contribution of this paper. The report establishes the technical baseline on which ADRES will continue building its analytical and system modeling capacities. For health system resource administrators in Latin America, it offers a reference case on the design decisions and operational obstacles that characterize health information centralization at national scale.

## Data Availability

The data underlying Figures 3 and 4 are available as Supporting Information files S1 and S2, submitted alongside this manuscript. The remaining operational metrics reported in this study are derived from ADRES's internal production environment and are not publicly available due to institutional data governance restrictions.

## Declarations

### Data availability

The reported operational metrics come from ADRES production systems and cannot be shared publicly due to institutional security restrictions. Requests for access to aggregated data for academic review purposes may be directed to the Directorate of Innovation, Analytics, and Knowledge Management of ADRES at.

### Ethical approval

This paper describes an analytical infrastructure and reports operational performance metrics; it does not conduct patient-level data analysis. The platform-enabled analyses that involve enrollment and service delivery data are carried out by the ADRES analytics team under the legal mandate established in Decree 228 of 2025 and Resolution 2275 of 2023, and are subject to applicable institutional governance frameworks. This paper does not constitute research on human subjects and does not require additional ethical approval.

### Author contributions

**D.A.G.J**.: Conceptualization, Data Curation, Investigation, Supervision, Writing — review and editing.

**D.B**.: Data Curation, Methodology, Software, Resources, Writing — review and editing.

**L.T.M.L**.: Data Curation, Validation, Writing — review and editing.

**D.C**.: Data Curation, Validation, Writing — review and editing.

**D.D.F**.: Software, Data Curation, Writing — review and editing.

**S.P**.: Methodology, Software, Data Curation, Writing — review and editing.

**F.L.M**.: Conceptualization, Funding Acquisition, Supervision, Writing — review and editing.

**J.C.B.P**.: Conceptualization, Investigation, Methodology, Project Administration, Writing — original draft, Writing — review and editing.

## Figure legends

**Figure 1**. Data volume by catalog — ADRES analytical platform, January–November 2025. Distribution of stored tables and records across the 21 production data catalogs. Highest-volume catalogs correspond to TUVA, sufficiency, BDUA, RIPS, and electronic billing. Cumulative production exceeding 1 trillion records. Source: Unity Catalog, ADRES, 2025.

**Figure 2**. Speed and concurrency — ADRES analytical platform, January–November 2025. Left: daily query count by functional group, with peak of 78,028 monthly queries in October 2025. Right: FEV-RIPS batch processing speed; 220 million invoices processed in three hours. Source: Databricks monitoring system, ADRES, 2025.

**Figure 3**. Computational capacity and query performance — ADRES analytical platform, 2025. Maximum deployed cluster sizing (up to 32 TB RAM, 4,096 vCPU). Response time on 100 billion records: ten seconds. Source: Databricks cluster configuration records, ADRES, 2025.

**Figure 4**. ADRES analytical platform architecture, 2025. Full technology stack organized by layers: on-premise sources, VPN/Azure connectivity, orchestration (ADF), distributed processing (Bronze/Silver/Gold layers in Databricks), governance (Unity Catalog, Azure Purview, DGD), and visualization (Power BI, Microsoft Fabric). Source: Directorate of Innovation, Analytics, and Knowledge Management, ADRES, 2025.

